# Estimation of Vaccine Efficacy for Variants that Emerge After the Placebo Group Is Vaccinated

**DOI:** 10.1101/2021.08.31.21262908

**Authors:** Dean Follmann, Michael Fay, Craig Magaret, Peter Gilbert

## Abstract

SARS-CoV-2 continues to evolve and the vaccine efficacy against variants is challenging to estimate. It is now common in phase III vaccine trials to provide vaccine to those randomized to placebo once efficacy has been demonstrated, precluding a direct assessment of placebo controlled vaccine efficacy after placebo vaccination. In this work we extend methods developed for estimating vaccine efficacy post placebo vaccination to allow variant specific time varying vaccine efficacy, where time is measured since vaccination. The key idea is to infer counterfactual strain specific placebo case counts by using surveillance data that provide the proportions of the different strains. This blending of clinical trial and observational data allows estimation of strain-specific time varying vaccine efficacy, or sieve effects, including for strains that emergent after placebo vaccination. The key requirements are that surveillance strain distribution accurately reflect the strain distribution for a placebo group, throughout follow-up after placebo group vaccination and that at least one strain is present before and after placebo vaccination. For illustration, we develop a Poisson approach for an idealized design under a rare disease assumption and then use a proportional hazards modeling to better reflect the complexities of field trials with staggered entry, crossover, and smoothly varying strain specific vaccine efficacy We evaluate these by theoretical work and simulations, and demonstrate that useful estimation of the efficacy profile is possible for strains that emerge after vaccination of the placebo group. An important principle is to incorporate sensitivity analyses to guard against mis-specfication of the strain distribution. We also provide an approach for use when genotyping of the infecting strains of the trial participants has not been done.

## 1 Introduction

Multiple COVID-19 vaccine trials have demonstrated substantial short term efficacy, though the durability of vaccination is, at this time, unknown. Recent work has developed methods to assess waning vaccine efficacy for vaccine trials where the placebo group is vaccinated [4], [12],[16], [3]. By assuming the effect of vaccination is the same over calendar time, the placebo-controlled vaccine efficacy profile is recoverable long after all placebo volunteers received vaccine. This work assumed that the vaccine efficacy, at any given time since vaccination, was similar for all circulating strains. However multiple new strains or variants have emerged with improved transmissibility and the vaccine efficacy for new strains may be less than for the original strains [11], [14].

In this paper, we develop approaches for settings where new virus strains emerge over time and the vaccine may have efficacy that varies by strain and time since vaccination. Differential vaccine efficacy by strain is known as sieve analysis [6], [7]. The simplest and most robust approach to sieve analysis is to apply the methods given above separately to each strain. However, this approach fails for strains that emerge after the placebo group is vaccinated and with little overlap of strains pre and post placebo vaccination, sieve effects may be poorly estimated. For such strains, new approaches are needed. In this work we develop a new method that leverages strain surveillance data [10]. It requires that the distribution of strains in the placebo trial participants (whether actual or counterfactual) matches the proportions observed in the community and that at least one strain is present before and after placebo vaccination. This allows imputation of strain specific case counts for a counterfactual placebo group which can recover placebo controlled vaccine efficacy, in a fashion similar to [4].

We extend this strain-specific analysis to a mark based approach, [8], [9], where the vaccine efficacy of a virus is assumed to vary smoothly with a quantitative phenotype of the virus, e.g. from a neutralization assay. These approaches allow for time-varying placebo controlled strain-specific vaccine efficacy profile to be recovered for stains that emerge after the placebo group is vaccinated.

We develop a basic Poisson approach for an idealized settings under a rare disease assumption and carefully delineate how bias is introduced if the surveillance strain distribution differs from the placebo group. Simulations are performed to illuminate key features. We then develop a Cox regression approach which seamlessly accommodates the complexities of phase III vaccine trials. The method is computationally intensive and can require up to *E* near copies of the dataset where *E* is the number of cases, though standard software can be used to fit the model. We also develop an approach that can be used if genotyping of viruses is not done within the trial, relying on variation in the strain distribution over time and space to modulate the overall vaccine efficacy. We evaluate the Cox approaches via simulation and analyze example datasets meant to approximate the Moderna and Pfizer Phase III efficacy trials over the period September 2020 to September 2021.

## 2 Surveillance Anchored Sieve Analysis

### 2.1 Preliminaries

We begin by reviewing the idealized development of [4]. Suppose we have a large placebo controlled vaccine trial with simultaneous entry and follow-up over a fixed period of time [0,T]. For simplicity we assume equal randomization to the two arms, this assumption is relaxed in the appendix. The trial is a deferred vaccination design and all placebo volunteers are vaccinated halfway through the trial. Thus period 1 [0,T/2) allows a contrast of vaccine versus placebo while period 2 [T/2,T] allows a contrast of immediate versus deferred vaccination. Let *A* = 0, 1 denote randomization to the placebo and vaccine, respectively, and define *X*_*AKS*_ as the total number of cases, i.e. first infections that result in disease, on arm *A* in period *K* who are infected with strain *S* = 0, 1, respectively wild-type and variant. We assume *X* is approximately Poisson which follows from a rare disease assumption. Define *E*(*X*_*AKS*_) = *ω*_*AKS*_ and let *θ*_0*KS*_ be the expected case count with strain=S in a placebo arm in period *K* whether actual (K=1) or counterfactual (K=2). Note that in period 1, *θ*_02*S*_ = *ω*_02*S*_ but in period 2 the placebo group is vaccinated so *θ*_01*S*_ ≠ *ω*_02*S*_ We define vaccine efficacy against strain S in period *K* as

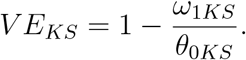

thus VE_1*S*_ is the the early effect of vaccine immediately after vaccination and VE_2*S*_ the late effect of vaccine after one period has elapsed. We can directly estimate the early vaccine efficacy as 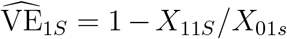, but in Period 2, we have no placebo group and *θ*_02*S*_ is not directly estimable.

Nonetheless, we can recover the placebo controlled late vaccine efficacy by assuming portability of the immediate effect of vaccination, as shown in [4]. Under this assumption *E*(*X*_02*S*_) = *ω*_02*S*_ = *θ*_02*S*_(1 *− V E*_1*S*_). Thus *θ*_02*S*_ = *ω*_02*S*_*/*(1 *− V E*_1*S*_) which is estimated by *X*_02*S*_*/*(*X*_01*S*_*/X*_11*S*_) We can estimate the late placebo controlled vaccine efficacy over period 2 as

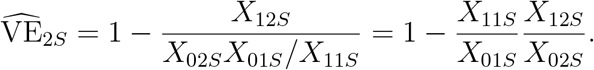

For a more elegant development involving Cox regression with smoothly varying VE that allows for staggered entry trials, dropout, covariate adjustment, etc. see [12],[16], [3]. This approach works fine if strains *S* = 0 and *S* = 1 circulate in both periods. But if *S* = 1 emerges in period 2 this approach will not work as *X*_011_ = 0.

### 2.2 New Strains After Placebo Vaccination

Suppose that in Period 1, only a wild-type strain *S* = 0 circulates, while in Period 2 a new variant strain, *S* = 1, emerges. We want to recover the early and late strain specific placebo controlled vaccine efficacies VE_1*S*_, VE_2*S*_, respectively, but have no placebo arm for *S* = 1. Suppose that at all the sites of the trial, community surveillance of the circulating strains is done. Let *p*_*KS*_ be the proportion of strain *S* = 0,1 in period *K* = 1, 2 in the community. Under the assumption that the distribution of infecting strains in the community, i.e. *p*_*KS*_, matches that of the placebo group in the trial, whether actual or counterfactual, we can recover time-varying placebo controlled vaccine efficacy. Note that since *p*_*K*0_ + *p*_*K*1_ = 1 the ratio *p*_*K*0_*/p*_*K*1_ determines *p*_*K*0_, *p*_*K*1_ so only the ratio need to be correctly specified.

Figure 1 provides a schematic of how this is possible. The key idea is to infer strain specific case counts for a Period 2 counterfactual placebo group. We do this in two stages. The early vaccine efficacy over Period 1 for *S* = 0 or wild type virus is estimated as 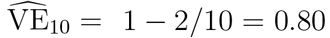. As shown above, in Period 2, this estimated 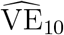 is applied to the newly vaccinated placebo arm, i.e. deferred vaccination arm, thus inferring 5 wild type cases. The community surveillance ratio of 2:1 for S=1:S=0, is then applied to the inferred 5 wild type cases, thus sequentially inferring 10 variant cases. With these counterfactual placebo cases for wild type and variant strains, placebo controlled variant vaccine efficacy for the newly vaccinated can be estimated as 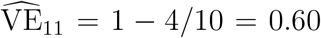. For those vaccinated one period earlier, the variant and wild type VEs are estimated as 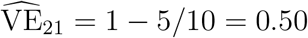 and 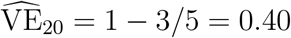, respectively.

**Figure 1:**
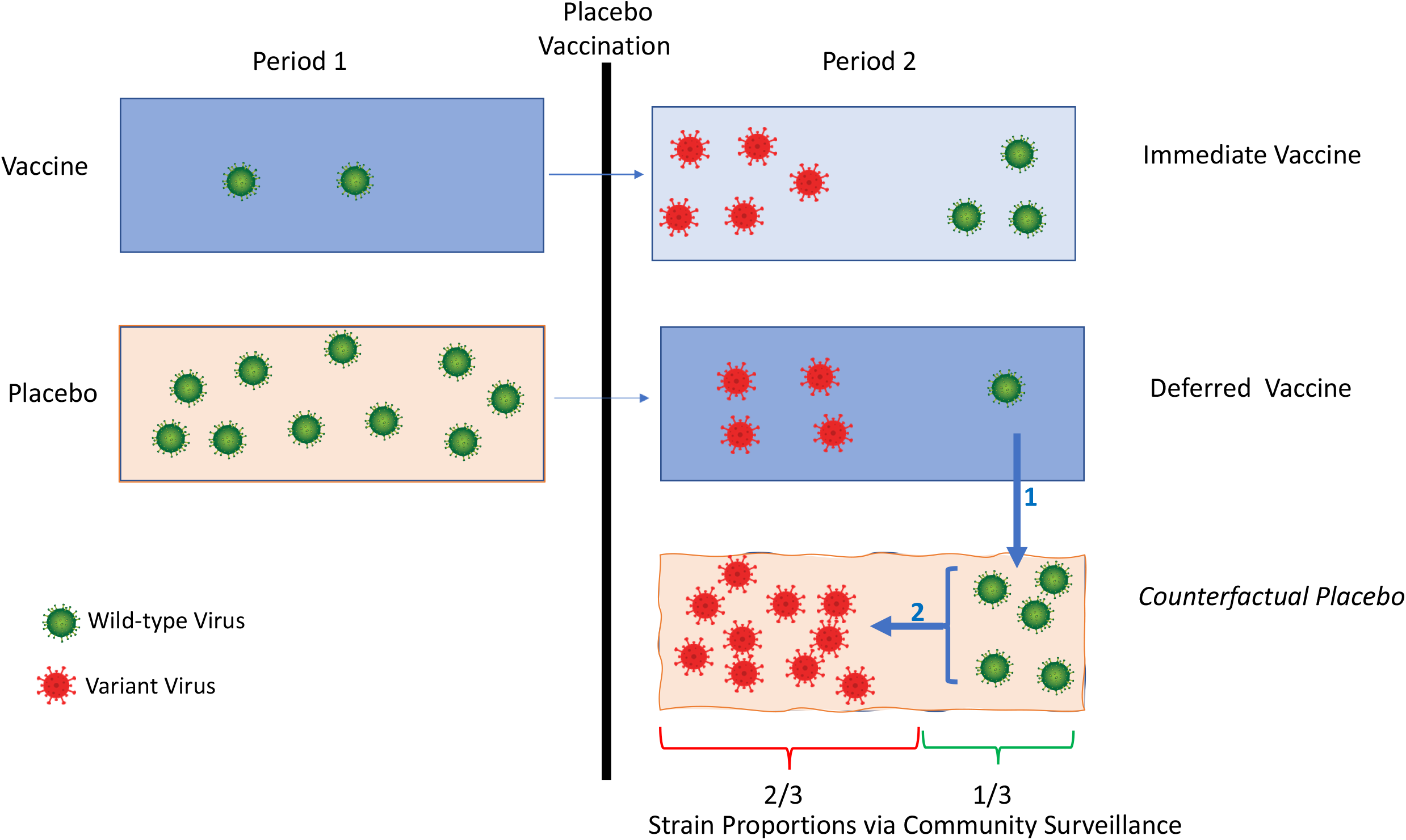
Imputation of strain specific cases for a counterfactual placebo group in period 2. Imputation #1 of 5 wild-type (green) cases follows from portability of wild type VE of 0.80 for the newly vaccinated which is estimated in period 1. Imputation # 2 of 10 variant (red) cases follows from the 2:1 variant:wild type case split seen in the surveillance cohort. With wild type and variant placebo case counts, early and late vaccine efficacy for wild type and variant strains is easy to calculate. For example, early vaccine efficay for the variant strain (VE_11_) is estimated as 1-4/10=0.60

Table 1 provides the relationship between the parameterization for the observed data from the trial, *E*(*X*_*AKS*_) = *ω*_*AKS*_, the surveillance based probabilities *p*_*KS*_, the overall expected placebo cases counts in period *K, θ*_*K*_, and VE_*KS*_. We call this approach surveillance anchored sieve analysis (SASA) as the surveillance proportions *p*_*KS*_ ’anchor’ or serve as fixed constants in the Poisson model. We assume they are based on a large sample relative to the trial and treat them as constants. With these as fixed constants, we can estimate the six parameters (*θ*_1_, *θ*_2_,VE_10_,VE_11_,VE_20_,VE_21_) using the six counts (*X*_010_, *X*_110_, *X*_020_, *X*_021_, *X*_120_, *X*_121_). For details see Appendix “GLM Analysis for the Poisson Model” which additionally allows Poisson error for the surveillance data.

**Table 1:** The relationship between mean case counts *ω*, Period 2 surveillance strain proportions *p*_20_, *p*_21_, overall placebo mean cases *θ*_*K*_, and time varying strain specific vaccine efficacy *V E*_*KS*_ for a two period deferred vaccination design. Only wild type *S* = 0 is observed in Period 1 but a new variant strain *S* = 1 emerges in Period 2.

| Group | Period 1 | Period 2 | Strain |
| --- | --- | --- | --- |
| Immediate Vaccination | $\omega_{110} = \theta_1(1 - VE_{10})$ | $\omega_{120} = \theta_2 p_{20}(1 - VE_{20})$ | S=0 Wild Type |
| Immediate Vaccination | – | $\omega_{121} = \theta_2 p_{21}(1 - VE_{21})$ | S=1 Variant |
| Deferred Vaccination | $\omega_{010} = \theta_1$ | $\omega_{020} = \theta_2 p_{20}(1 - VE_{10})$ | S=0 Wild Type |
| Deferred Vaccination | – | $\omega_{021} = \theta_2 p_{21}(1 - VE_{11})$ | S=1 Variant |

### 2.3 Bias Evaluation

The key assumption of SASA is that the surveillance distribution of strains matches what we would see in an unvaccinated placebo group, whether actual or counterfactual. The matching can be directly examined during the placebo controlled era of the trial. In the post placebo vaccination era, it is an unexaminable assumption. We next explore the potential bias in VE_1_ from a surveillance system that is somewhat different from a counterfactual placebo group.

The expected ratio of cases for S=0 to S=1 in Period 2 for the recently vaccinated placebo group is

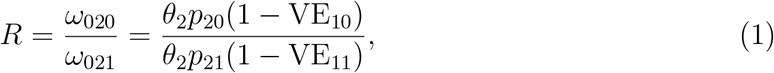

which can be solved for VE_11_, the VE for the variant strain *S* = 1 in the newly vaccinated:

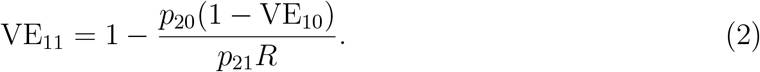

To estimate VE_11_ we can plug in *p*_20_, *p*_21_ from the surveillance distribution, VE_10_ estimated from period 1, and *R* estimated from Period 2 data into (2). Suppose that the surveillance proportions are incorrectly specified as 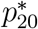 and 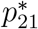. Then the RHS of (2) is not VE_11_ but 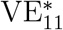

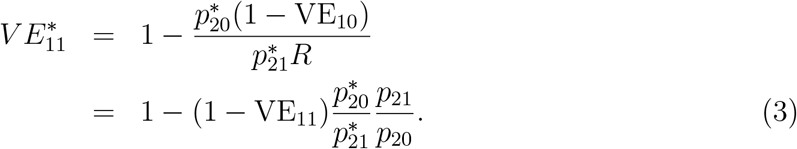

where the second line follows by replacing *R* with the RHS of (1). An analogous result obtains for VE_21_, the late vaccine efficacy for the new strain following 1 period of vaccination, with VE_21_ replacing VE_11_ in equation (3).

We can evaluate (3) to explore the extent of the bias. For example if the truth is VE_11_=0.90 and *p*_21_ = 0.50 but we use 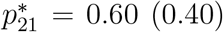 then 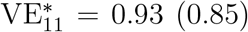 both of which are within 6% of the true value. On the other hand, if VE_11_ = 0.50 then the 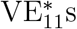 are 0.67 (0.25) which are substantially biased. The potential bias increases with decreasing VE_11_. Another feature is that bias can be substantial for rare variants. Suppose VE_11_=0.70 with true *p*_21_ = .05 but we use 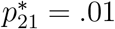 Then 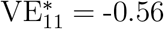. Thus in applying these methods one should be cautious about rare variants.

For the wild-type strain, there is no bias for VE_20_ with mis-specification of the *p*s as the proportion 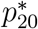 cancels out of the ratio

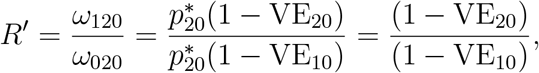

and we obtain VE_20_ by plugging in VE_10_ and *R′*

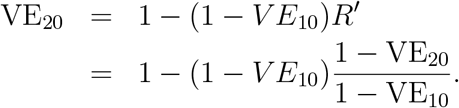

Effectively, the early and late VEs of the wild-type strain are estimated by restricting to wild-type data in periods 1 and 2, thus durability of vaccine efficacy, i.e. VE_10_= VE_20_ does not depend on the *p*_*ks*_s.

### 2.4 Simulation

To evaluate the statistical performance of the SASA estimates of vaccine efficacy we conducted a small simulation meant to loosely track the simplified version of the Pfizer or Moderna Phase III Trials of mRNA vaccines. To generate the data, we set the placebo mean count as *θ*_1_ = *θ*_10_ + *θ*_11_=1000 with an early vaccine efficacy of 90% for wild type virus. During period 2, a new variant emerges and accounts for half of the infections as determined by surveillance sampling. This is meant to approximate the emergence of a new variant in the US with an expectation it will dominate in the summer (see appendix). We set the counterfactual placebo group mean case count as *θ*_2_ = *θ*_20_ + *θ*_21_=500. We consider five scenarios, the last two where the strain distribution is mis-specified.

1. Vaccine efficacy is 90% throughout follow-up for wild type and variant strains with *p*_20_, *p*_21_ = 0.50, 0.50
2. Vaccine efficacy wanes from 90% to 80% for wildtype and from 80% to 60% for variant with *p*_20_, *p*_21_ = 0.50, 0.50.
3. Vaccine efficacy is 90% throughout follow-up for wild type and variant strains with *p*_20_, *p*_21_ = 0.20, 0.80
4. Vaccine efficacy is 90% throughout follow-up for wild type and variant strains with *p*_20_, *p*_21_ = 0.50, 0.50, but incorrect offsets of 0.60,0.40 are used.
5. Vaccine efficacy is 90% throughout follow-up for wild type and variant strains with *p*_20_, *p*_21_ = 0.50, 0.50, but incorrect offsets of 0.40,0.60 are used.

We estimate *θ*_1_, *θ*_2_ and the VE parameters using Poisson regression with parameterization given by Table 1 and correct (Cases 1-3) or biased (Cases 4-5) offsets. Code is provided in the appendix and results in Table 2 which demonstrates that we can accurately recover wild type VEs for period 1 and period 2 for all scenarios. We recover accurately recover the variant VE under Cases 1-3 and with moderate bias for Cases 4-5.

**Table 2:** Simulated performance of Surveillance Anchored Sieve Analysis for an idealized two period Deferred Vaccination Design. During Period 1 (Period 2) the expected actual (counterfactual) placebo case count is 1,000 (500). The true VE_*KS*_ = 0.90 except for scenario 2 with VE_20_, VE_21_, = 0.80,0.60. The VE columns report the Monte Carlo means and (standard deviations). The right columns report the rejection rates for 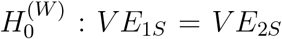 and 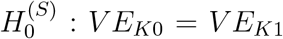, respectively, based on a Wald test. 10,000 trials are simulated per row.

| Scenario | $VE_{10}$ | $VE_{20}$ | $VE_{11}$ | $VE_{21}$ | VE change? | | Sieve Effect? | |
| --- | --- | --- | --- | --- | --- | --- | --- | --- |
| | | | | | $S = 0$ | $S = 1$ | Period 1 | Period 2 |
| 1- Reference | 0.8998<br>(0.0105) | 0.8955<br>(0.0336) | 0.8955<br>(0.0337) | 0.8958<br>(0.0337) | 0.0499 | 0.0451 | 0.0475 | 0.0460 |
| 2- Waning VE | 0.9000<br>(0.0105) | 0.7909<br>(0.0598) | 0.7902<br>(0.0609) | 0.5814<br>(0.1143) | 0.8235 | 0.9856 | 0.8055 | 0.9709 |
| 3- $p_{20} = 0.80$ | 0.8999<br>(0.0105) | 0.8868<br>(0.0626) | 0.8868<br>(0.0540) | 0.8870<br>(0.0546) | 0.0415 | 0.0496 | 0.0373 | 0.0438 |
| 4- $p_{20}/p_{21} = 1.50^*$ | 0.8998<br>(0.0104) | 0.8954<br>(0.0332) | 0.8429<br>(0.0519) | 0.8425<br>(0.0513) | 0.0469 | 0.0477 | 0.2856 | 0.2840 |
| 5- $p_{20}/p_{21} = 0.67^*$ | 0.9000<br>(0.0105) | 0.8952<br>(0.0336) | 0.9298<br>(0.0230) | 0.9298<br>(0.0228) | 0.0454 | 0.0470 | 0.2793 | 0.2804 |

\* True ratio is as given but offsets incorrectly use ratio of 1.00

Under case 1, the ratio of variances for estimates that are indirectly estimated (VE_20_, VE_21_,VE_11_) compared to the directly estimated VE_10_ is about (.033*/*.01)^2^ *≈* 11. This is partly due to having about 4 times more expected placebo cases in period 1 compared to period 2, and partly due to VE_10_ being directly estimated. Nonetheless, a typical confidence interval for an indirectly estimated VE is still fairly tight at about (0.84, 0.96). The probability of rejecting at *α* = 0.05 no waning efficacy (i.e., no change in early vs. late efficacy), 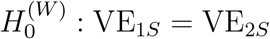, or rejecting no sieve effect, 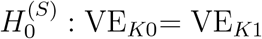, for *K* = 1,2, are appropriately about 0.05 under their respective nulls.

Under case 2 with waning VE, the estimates are again unbiased, and with greater standard deviations, due to the VEs being farther from 1.00. Power to detect waning efficacy is high for wild type (a contrast of 4 random variables) and very high for variant (a contrast of 2 random variables). Power to a sieve effect matches the powers for waning efficacy under case 2 because the expected Wald statistics for strain-specific waning efficacy equals the expected Wald statistics for period specific sieve effects.

Under scenario 3 with the variant dominant in period 2, the estimates again appear unbiased but with an increase in the standard deviation by about 50% compared to reference case 1 for the indirectly estimated parameters. The standard deviation for VE_11_ is unchanged.

Under scenarios 4 and 5 with mis-specification of the proportions in period 2, the wild type estimates are unbiased and there is no inflation of the type I error rate for testing of waning efficacy for the wild type or variant strains. There is substantial inflation of the type I error rate for a test of sieve effect, but moderate bias of about -7% and +3% respectively, for the VE of the variant strain. The inflation of the type I error rate for a test of sieving is noticeable.

The Poisson approach is useful for conceptual understanding and high-level evaluation of an idealized situation with a rare disease. However, this approach does not easily accommodate non-constant strain specific vaccine efficacy that can vary smoothly across the pre and post crossover eras. Cox modeling handles this easily and is discussed next.

## 3 Cox Models

The Cox model can seamlessly accommodate staggered entry, dropout, covariate adjustment, placebo attack rate varying with calendar time, stratification by site, strains coming and going, and strain specific VE varying smoothly with time. The Cox model can recover the per-exposure vaccine efficacy provided 1) the exposures to each infecting strain are the same in the placebo and vaccine arm, which should be achieved in a blinded trial which is a small portion of the population and 2) the per-exposure disease probabilities are correctly specified (e.g. as a function of time or covariates) [15], [6], [5]. The main disadvantage of the Cox approach is computational complexity.

Following the parameterization of [3], let *t* be the time since study initiation, 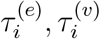, be the enrollment and vaccination times for person *i*, and *Z*_*i*_(*t*) be the indicator of whether person *i* is vaccinated at time *t*. As an example parameterization that allows for a smoothly declining vaccine efficacy, suppose that the hazard for the risk of first infection by stain *S* = *s* is given by

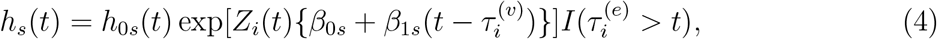

where *h*_0*s*_(*t*) is the placebo baseline hazard (whether actual or counterfactual) for infection by strain *S*, and *I*() is the indicator function. This formulation (4) reduces to the Prentice approach to competing risks applied to each strain separately. However, vaccine efficacy for strains that emerge after the placebo group is vaccinated cannot be estimated with this approach as *Z*(*t*) = 1 for everyone after placebo vaccination. To see this, note that the partial likelihood contribution for a strain *S* = 4 case at calendar time *t* is

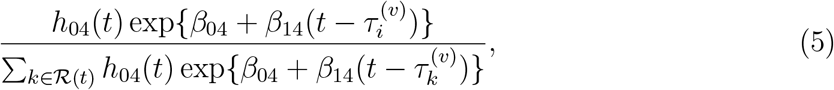

where *ℛ*(*t*) is the set of volunteers who are still at risk at time *t*. Because *S* = 4 emerges after placebo vaccination, *β*_04_ cancels out for all partial likelihood contributions, so *β*_04_ and VE_4_(*t*) = 1 *−* exp(*β*_04_ + *β*_14_*t*) are not estimable. However, *β*_14_ and thus whether VE_4_(*s*) is constant, can be assessed. We call this the strain specific sieve approach (SSSA).

For Cox regression, the key idea of Figure 1 is rendered as an assumption that the cause-specific hazards have a relationship anchored by the strain distribution of the surveillance data. Let *P* (*t, s*) be the true proportion of strains *S* viruses at time *t* in the surveillance cohort, and let *h*_0_(*t*) be the hazard of infection by any strain at calendar time *t*. We write

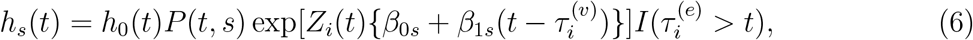

where we have substituted *h*_0*S*_(*t*) from (4) with *h*_0_(*t*)*P* (*t, s*), a nuisance parameter times a known function.

To illustrate how the estimation works, consider the partial likelihood contribution for a person *i* who is infected with strain *S* = 4 at calendar time *t* after all have been vaccinated. The likelihood contribution is

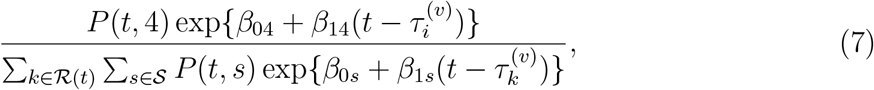

where *𝒮* is the set of circulating strains observed over followup. Note that *β*_04_ remains in the likelihood throughout follow-up. So even if strain S=4 emerges after all have been vaccinated, we recover the immediate post vaccination vaccine efficacy for S=4 as 1 *−* exp(*β*_04_). This recovery is analogous to what we saw in Figure 1 which demonstrated recovery of Period 1 VE for a strain that emerged in Period 2.

We can also apply SASA to the setting where we have a phenotypic mark *V* (*s*) for strain *s*, and vaccine efficacy depends on *V* (*s*). This builds on work by [8], [9], and is illustrated for HIV by [1]. To fix ideas, suppose that *V* (*s*) is the readout from a neutralization assay using pseudo-virus *s* with pooled sera collected shortly after vaccination. An example mark-based parameterization with time constant vaccine efficacy is given by

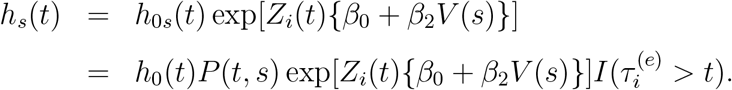

With this model we have the vaccine efficacy vary smoothly with the mark but constant in time as

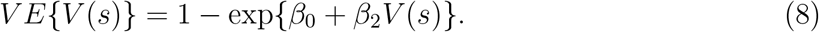

Due to the potential for bias due to mis-specification of *P* (*t, s*) it is appealing to apply the mark based approach with *h*_0*s*_(*t*), unspecified. For strains that are present in the precrossover era, *h*_0*s*_(*t*) can be left unspecified. Indeed each infecting virus can be viewed as a different strain so that there are as many strains as cases. Effectively, each strain defines a stratum in the Cox model. However for strains that emerge post-crossover, there is a subtly. Post crossover data provides no information about *β*_0_ or *β*_2_. To see this note that in the post-crossover era the partial likelihood contribution reduces to a constant

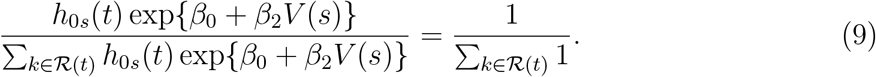

As a consequence of this subtly, suppose there are two strains with different marks V(0),V(1) in the pre-crossover era, and a new strain with yet another V(2) mark in the postcrossover era. One can estimate (8) with *h*_0*S*_(*t*) unspecified, using a data set with all the pre and post crossover data, but the post-crossover data is not used and it is a bald extrapolation to plug in V(2) into (8) to estimate VE for the emergent strain.

We note that one can view (8) with *h*_0*s*_(*t*) unspecified as a kind of smoothly parameterized Method B Lunn-McNeil approach to competing risks [13]. Method B of Lunn McNeil with two strains can be written as

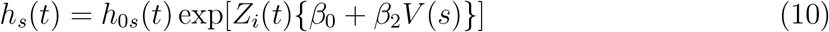

where *V* (*s*) = *s* for *s* = 0, 1.

## 4 Estimation

Estimation of the SASA Cox model is non-standard due to the presence of *P* (*t, s*), which is estimated by surveillance data, and because there can be many different variants. In this section, we assume that the idealized continuous function of time *P* (*t, s*) has been estimated over periods and use the notation *p*_*L*_(*k, s*) which is piecewise constant for period *k*, say two weeks, and location *L*.

### 4.1 Specification of *p*_*L*_(*k, s*)

We estimate *p*_*L*_(*k, s*) using the GISAID database (https://www.gisaid.org), which contains the genetic sequences of infecting SARS-CoV-2 strains from around the globe starting in January 2020 [2]. The sequences used for this paper are from the corpus of whole-genome sequences made available be GISAID, and they were downloaded on 08 July 2021. All sequences in this corpus from the United States (n = 539,812) and from Mexico (n = 13,557). The GISAID accession numbers for the sequences used in this study are available in the supplementary materials. We determine their lineages and WHO variant status using the Phylogenetic Assignment of Named Global Outbreak LINeages (PANGOLIN) (https://github.com/cov-lineages/pangolin) software. Additionally, each sequence has information about its country of origin and the date of submission, and for the United States, the state of the submitting lab is also included.

Specification of *p*_*L*_(*k, s*) depends on the granularity needed for analysis, the abundance of the different strains, the similarity of the strain distribution over location, and assumptions or data about which variants/lineages have similar vaccine efficacy over time. A coarse analysis might be to group lineages/variants into two or three strains, say Delta, Alpha, and otherwise. Or a set of 4 or 5 variants might be initially examined and then pooled into a smaller number of strains with similar vaccine efficacy. The location *L* could be regional and *p*_*L*_(*k, s*) would apply to all clinical trial sites in the region. If *p*_*L*_(*k, s*) is close to 0 and a strain *s* case occurs in week *k* at location *L*, it will have substantial influence. We thus recommend not counting cases for which *p*_*L*_(*k, s*) *>* 0.10.

A mark-based analysis is more complex as in principle each infecting virus might yield a unique mark *V* (*s*). If each infecting virus were classified as a different strain then *p*_*L*_(*k, s*) is zero except the one *k* when a case of type *s* occurs and *p*_*L*_(*k, s*) becomes 1. Such a strain distribution implies that post-crossover data provide no information about time-constant sieve effects. Thus to glean information post-crossover, we are led to defining *p*_*L*_(*k, s*) as fairly coarse as described above. Figure 2 shows the weekly distribution of WHO lineages over time for 10 regions of the contiguous United States from January 2020 through June 2021.

**Figure 2:**
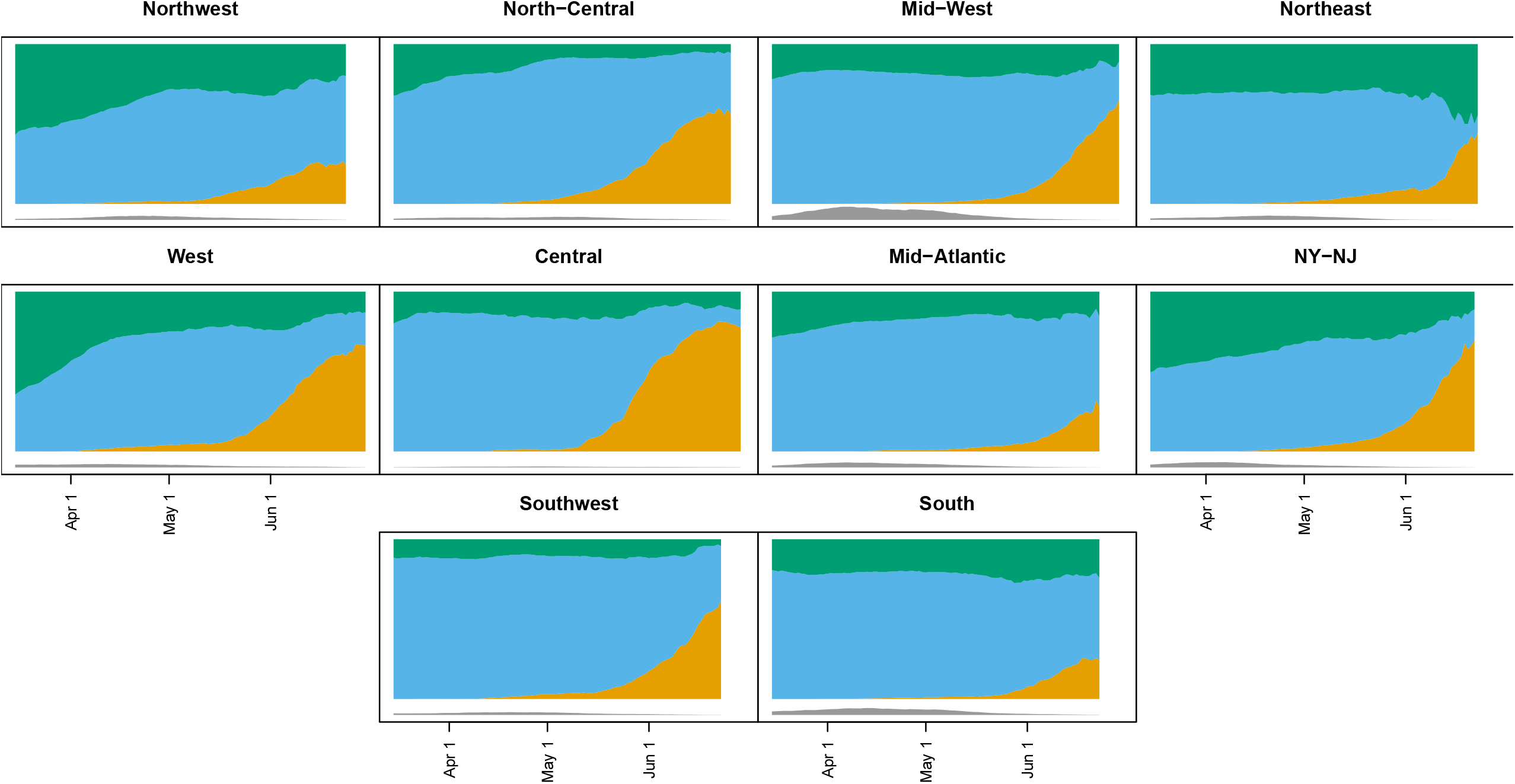
The plots of the moving averages (+/- 7 days) of proportions of different variants out of those not missing variant information. The Delta variant (including Delta+K417N) is orange, the Alpha variant is blue, and all other non-missing variants are green. Values are plotted from March 15, 2021 to June 30, 2021, but some regions do not have data all the way until June 30 (e.g., Northwest). Underneath the main figure are gray regions which are proportional to the sample size of the averages, with the bottom of the color plot at the value n=20,000. For example, the largest sample size is 16,469 for the Mid-West region on April 8, 2021, but most have much smaller sample sizes especially in late June. Regions defined at https://fortune.com/2021/07/07/the-delta-variant-is-now-dominant-in-the-u-s-see-the-states-where-its-most-prevalent

### 4.2 Estimation of Cox Model Parameters

The partial likelihood is the product of contributions like that of (7). For each two-week period we create a dataset of all volunteers who have not had an event before that period. For each event within the two-week period we determine each individual’s vaccination status and time since vaccination at the time of that event. For a dataset of *M* strains, we make *M* copies of the information for each person for period *k* but with an offset which equals log*{p*(*k, s*)*}* for strain *s* and an indicator of an event. Once an individual has an event within period *k* she is removed from the risk set for that period and subsequent periods.

So for a study with 100 periods and 4 strains, we make 400 strata each of which includes approximately *N* individuals, where *N* is the number randomized. If we further stratify sites by say 4 regions within which *P* (*k, s*) is homogeneous this increases the size of the dataset 4-fold to approximately 2000. Details with a simple example dataset and SAS code are provided in the appendix.

## 5 Simulation

In this section we apply the Cox model approach to a scenario that loosely approximates the Moderna and Pfizer vaccine trials. The trial enrolls 30,000 subjects uniformly in September and randomizes equally to vaccine or placebo. The placebo attack rate is constant from 1 September 2020 through 31 March 2021 with a mean number of placebo cases of about 1000. All placebo volunteers receive vaccine on 1 April 2021 and from April to 1 September 2021, the mean counterfactual placebo case count is about 750. There are three strains S=0,1,2 with strains 1 and 2 emerging in February and June respectively, and strain 0 disappearing in July (see appendix). We define a period as one month and specify a mark *V* (*s*) = s for *s* = 0, 1, 2. We generate data under two scenarios using a mark model with time constant vaccine efficacy

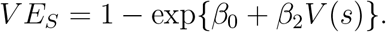

Under scenario 1) (*β*_0_, *β*_1_) = (log(.1), 0) so VE_0_,VE_1_,VE_2_ = (0.90,0.90,0.90) while under scenario 2) (*β*_0_, *β*_1_) = (*−* 2.5, 1.0) so VE_0_,VE_1_,VE_2_ = (0.92, 0.78, 0.39). We estimate 3 different models with time-constant VE.

- Strain Specific Sieve Analysis (SSSA) VEs for each strain.

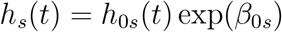

- SASA with separate VEs for each strain

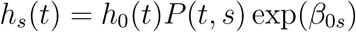

- Mark based SASA where time constant VE varies smoothly with each strain

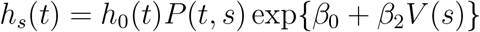

To examine durability of vaccine efficacy we additionally fit (6) to the first two (non-mark) models. This adds a strain-specific log-linear term *β*_1*s*_(*t − τ* ^(*v*)^) so the hazard is proportional to exp*{β*_0*s*_ + *β*_1*s*_(*t − τ* ^(*v*)^)*}* To illustrate a sensitivity analyses we fit models where the most common strain in any given month is specified as 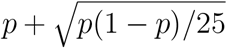 instead of *p* while the other strains are proportionally reduced so that the remaining *p*(*t, s*) sums to one. We analogously reduce the most common strain by 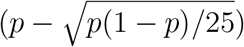. Note that for *p*_0_ = 0.50 with two strains perturbs to *p*_0_ = 0.60, *p*_1_ = 0.40 or *p*_0_ = 0.40, *p*_1_ = 0.60 respectively as was done in the Poisson simulations. We denote these perturbations by * and **, respectively.

The fitted estimates for vaccine efficacy from the constant VE models are given in the left half of Table 3 while the right half provides the slope estimates from the time varying VE models. The top half corresponds to scenario 1 with constant 90% VE for all strains. The first 3 pairs of rows are for correctly specified models. All approaches are unbiased for all VE estimates and all methods have similar standard deviations for VE_0_. For VE_1_ SASA-Mark is more efficient than SASA which is more efficient than SSSA with substantial efficiency gains (ratio of squared standard deviations) of (.0245*/*.0138)^2^ = 3.15 for SASA Mark compared to SSSA. VE_2_ is not estimable under the unspecified approach which has the same estimated standard deviation for SASA and SASA Mark. Both SSSA and SASA provide unbiased estimates of *β*_1*S*_ = 0.

**Table 3:** Monte Carlo estimates and (standard deviations) of vaccine efficacy for 100 simulated deferred vaccination trials. Estimates are provided both for strain specific and mark parameterized vaccine efficacy models. Two scenarios are evaluated with VE_0_, VE_1_, VE_2_ = (0.90, 0.90, 0.90) for the top half and with VE_0_, VE_1_, VE_2_ = (0.92,0.78,0.39) for the bottom half. VE is constant over time. The left half fits models with a time constant VE, while the right half fits models with a log-linear decline in VE. The unstarred rows denote correct specification of *p*(*k, s*) while * (**) denote mis-specifications.

|  | Constant VE |  |  | Time-Varying VE |  |  |
| --- | --- | --- | --- | --- | --- | --- |
| | $VE_0$ | $VE_1$ | $VE_2$ | $\beta_{10}$ | $\beta_{11}$ | $\beta_{21}$ |
| TRUTH | 0.90 | 0.90 | 0.90 | 0.00 | 0.00 | 0.00 |
| SSSA | 0.901<br>(.0099) | 0.899<br>(0.0245) | -<br>- | 0.007<br>(0.0431) | -0.001<br>(0.0268) | -0.004<br>(0.0533) |
| SASA | 0.901<br>(.0093) | 0.900<br>(0.0176) | 0.900<br>(0.0250) | 0.006<br>(0.0403) | -0.001<br>(0.0274) | -0.002<br>(0.0516) |
| SASA Mark | 0.901<br>(0.0091) | 0.901<br>(0.0138) | 0.899<br>(0.0250) |  |  |  |
| SASA* | 0.904<br>(.0091) | 0.879<br>(0.0228) | 0.877<br>(0.0287) | -0.011<br>(0.0405) | -0.008<br>(0.0272) | 0.016<br>(0.0518) |
| SASA Mark* | 0.902<br>(0.0089) | 0.891<br>(0.0147) | 0.876<br>(0.0288) |  |  |  |
| SASA** | 0.904<br>(.0094) | 0.882<br>(0.0171) | 0.920<br>(0.0167) | -0.004<br>(0.0374) | -0.010<br>(0.0287) | 0.011<br>(0.0516) |
| SASA Mark** | 0.899<br>(0.0093) | 0.908<br>(0.0112) | 0.915<br>(0.0174) |  |  |  |
| TRUTH | 0.92 | 0.78 | 0.39 | 0.00 | 0.00 | 0.00 |
| SSSA | 0.918<br>(.0098) | 0.776<br>(0.0436) | -<br>- | -0.000<br>(0.0504) | -0.001<br>(0.0194) | -0.007<br>(0.0194) |
| SASA | 0.917<br>(.0090) | 0.778<br>(0.0296) | 0.390<br>(0.1039) | -0.003<br>(0.0468) | -0.000<br>(0.0193) | -0.002<br>(0.0181) |
| SASA Mark | 0.917<br>(0.0079) | 0.776<br>(0.0234) | 0.387<br>(0.1034) |  |  |  |
| SASA* | 0.919<br>(.0088) | 0.738<br>(0.0376) | 0.271<br>(0.1256) | -0.0127<br>(0.0482) | -0.0146<br>(0.0190) | 0.010<br>(0.0183) |
| SASA Mark* | 0.916<br>(0.0080) | 0.758<br>(0.0260) | 0.300<br>(0.1211) |  |  |  |
| SASA** | 0.922<br>(.0088) | 0.774<br>(0.0249) | 0.596<br>(0.0584) | -.015<br>(0.0412) | -0.014<br>(0.0196) | 0.008<br>(0.0183) |
| SASA Mark** | 0.914<br>(0.0082) | 0.821<br>(0.0183) | 0.626<br>(0.0583) |  |  |  |

Looking at then next 4 pairs of rows we see that the SASA results are fairly robust to mis-specification of the strain distribution with mean VE_*S*_s estimates ranging from 0.88 to 0.92 for SASA and SASA-Mark. The estimates of the slope are robust to mis-specification of *p*_*L*_(*k, s*) and their standard deviations are very similar to when *p*_*L*_(*k, s*) is correctly specified.

The bottom panel reports results for scenario 2 with a strong sieve effect. Under correct specification (first 3 pairs of rows) all methods appear unbiased for VE though with larger standard deviations for 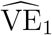 and 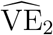 compared to VE_1_,VE_2_ = (0.90, 0.90). The efficiency gain for VE_1_ under SASA Mark is (.0234*/*.0436)^2^ = 3.47 compared to SSSA which is similar to the setting with VE_1_ = 0.90. Under mis-specification we see modest bias for VE_1_ which ranges from (0.74 to 0.82) for the various approaches, and more substantial bias for VE_2_ which ranges from 0.27 to 0.63 for the various approaches. As before, all approaches appear unbiased for *β*_1*S*_ so vaccine durability is accurately assessed.

In summary, the SSSA approach, which treats each strain completely separately and is free of concerns about *p*(*k, s*) mis-specification, is the most robust approach for estimating VE_*S*_ and also provides reliable inference about strain specific waning or *β*_1*S*_. For strains that are unreliably estimated with SSSA, or emerge post crossover, SASA can add useful information. For high VE, SASA is fairly robust while for low VE it is less so. Nonetheless, even for strains with lower VE, the analysis is informative. In scenario 2 we could conclude that strain 2 has lower efficacy than strain 0 and non-zero efficacy. Such conclusions are not possible without employing SASA. Use of the mark parameterized approach can result in greater efficiency in some scenarios with just 3 strains. With a large number of strains, a mark based approach should be even more efficient.

In the appendix we provide additional simulations with a much lower attack rate with an expected number of placebo cases in the pre-crossover and post-crossover eras of 130 and 100, respectively. This lower attack rate is meant to roughly approximate the Novavax phase III clinical trial. The behavior of the different methods are relatively similar, though the standard deviations are larger.

In practice we recommend reporting a union of confidence intervals for different perturbations of the *p*(*k, s*)s, to properly convey uncertainty. The specific perturbation would depend on the trial trial being analyzed. Because of the potential for rare variants to have substantial impact Sensitivity analyses such as rerunning the model after removing cases in turn, can be useful to see if some cases are overly influential. In our development, we used a linear function of time to describe waning vaccine efficacy In practice splines or polynomials may provide a better fit and should be evaluated.

## 6 Missing Strain Information

In practice, genotyping of the infecting strains within a trial takes time so the analysis outlined above might lag several months. Additionally, not all sequences can be genotyped, especially for those with higher cycle thresholds (i.e. smaller amount of virus). In such cases one can leverage the variation in the strain distribution over time and space to look for time-varying sieve. Suppose that at a generic location the SASA model holds

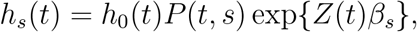

but we do not have the infecting stain for the cases. Thus the overall hazard is approximately

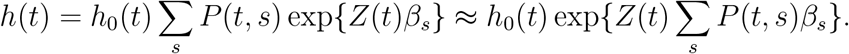

The approximation is exact if *β*_*s*_ = *β*, or *P*(*t, s*) = 1 for any *s*. We can fit the RHS using standard software, stratifying by location, and test for a (time-constant) sieve effect. Importantly, post-crossover data is not used in this approach as the covariates *P*(*t*, 0), *P*(*t*, 1), …, *P*(*t, M*) are identical for all subjects post-crossover.

A time-varying sieve effect can also be assessed. If the true strain specific hazard is

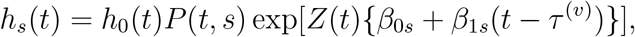

then the overall hazard is approximately

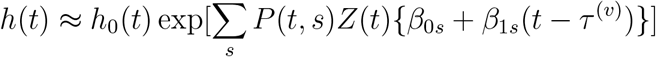

Post crossover, *β*_0*s*_ cancels out of the partial likelihood as all have *Z*(*t*) = 1, however *β*_1*s*_ does not. Thus we can test whether efficacy wanes differentially for different strains, including those at emerge post-crossover.

To explore the behavior of this approximation, we re-ran the simulation setup of the previous section with correct specification of *p*(*k, s*). The results are provided in Table 4. We see that the estimates of VE_*S*_ appear unbiased except for VE_1_ in the second scenario, however, it has large uncertainty and is within one standard deviation of the truth. The estimates of *β*_1*s*_ all appear unbiased. Unsurprisingly, there is some loss of efficiency compared to the strain specific sieve analysis which has genotypes the infecting strains. For VE_1_ there is little loss in efficiency. For VE_2_ = 0.90 the ratio of Monte Carlo variances (approximate/SSSA) is (.0496*/*.0245)^2^ *≈* 4.0 while for *β*_11_ the ratio is (0.0736*/*0.0268)^2^ *≈* 7.5. Thus there is an appreciable loss of efficiency here. Nonetheless useful estimation is still possible with this simple approximation.

**Table 4:**
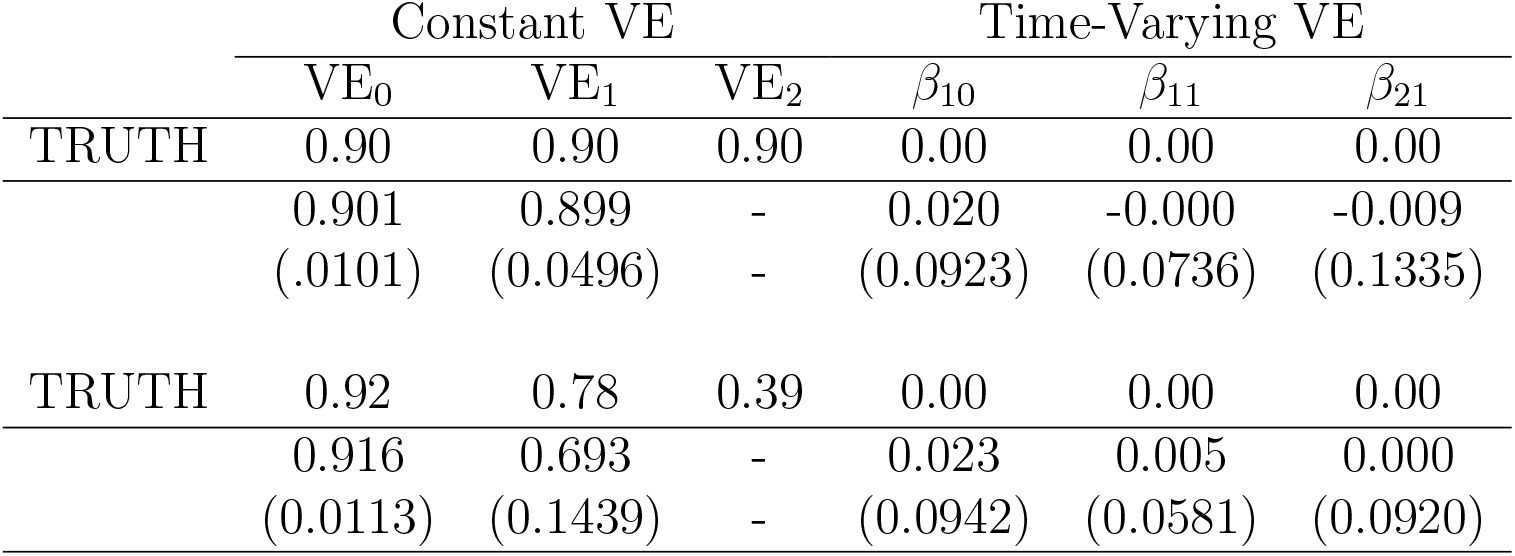
Monte Carlo estimates and (standard deviations) of vaccine efficacy for 100 simulated deferred vaccination trials. Estimation is based on data with missing strain information and an approximate hazard that treats *p*(*k, s*) as a covariate. Two scenarios are evaluated with (VE_0_, VE_1_, VE_2_) = (0.90,0.90, 0.90) for the top half and with (VE_0_, VE_1_, VE_2_) = (0.92,0.78,0.39) for the bottom half. VE is constant over time. The left half fits models with a time constant VE, while the right half fits models with a log-linear decline in VE.

|  | Constant VE |  |  | Time-Varying VE |  |  |
| --- | --- | --- | --- | --- | --- | --- |
| | $VE_0$ | $VE_1$ | $VE_2$ | $\beta_{10}$ | $\beta_{11}$ | $\beta_{21}$ |
| TRUTH | 0.90 | 0.90 | 0.90 | 0.00 | 0.00 | 0.00 |
|  | 0.901 | 0.899 | - | 0.020 | -0.000 | -0.009 |
|  | (.0101) | (0.0496) | - | (0.0923) | (0.0736) | (0.1335) |
| TRUTH | 0.92 | 0.78 | 0.39 | 0.00 | 0.00 | 0.00 |
|  | 0.916 | 0.693 | - | 0.023 | 0.005 | 0.000 |
|  | (0.0113) | (0.1439) | - | (0.0942) | (0.0581) | (0.0920) |

**Table 5:**
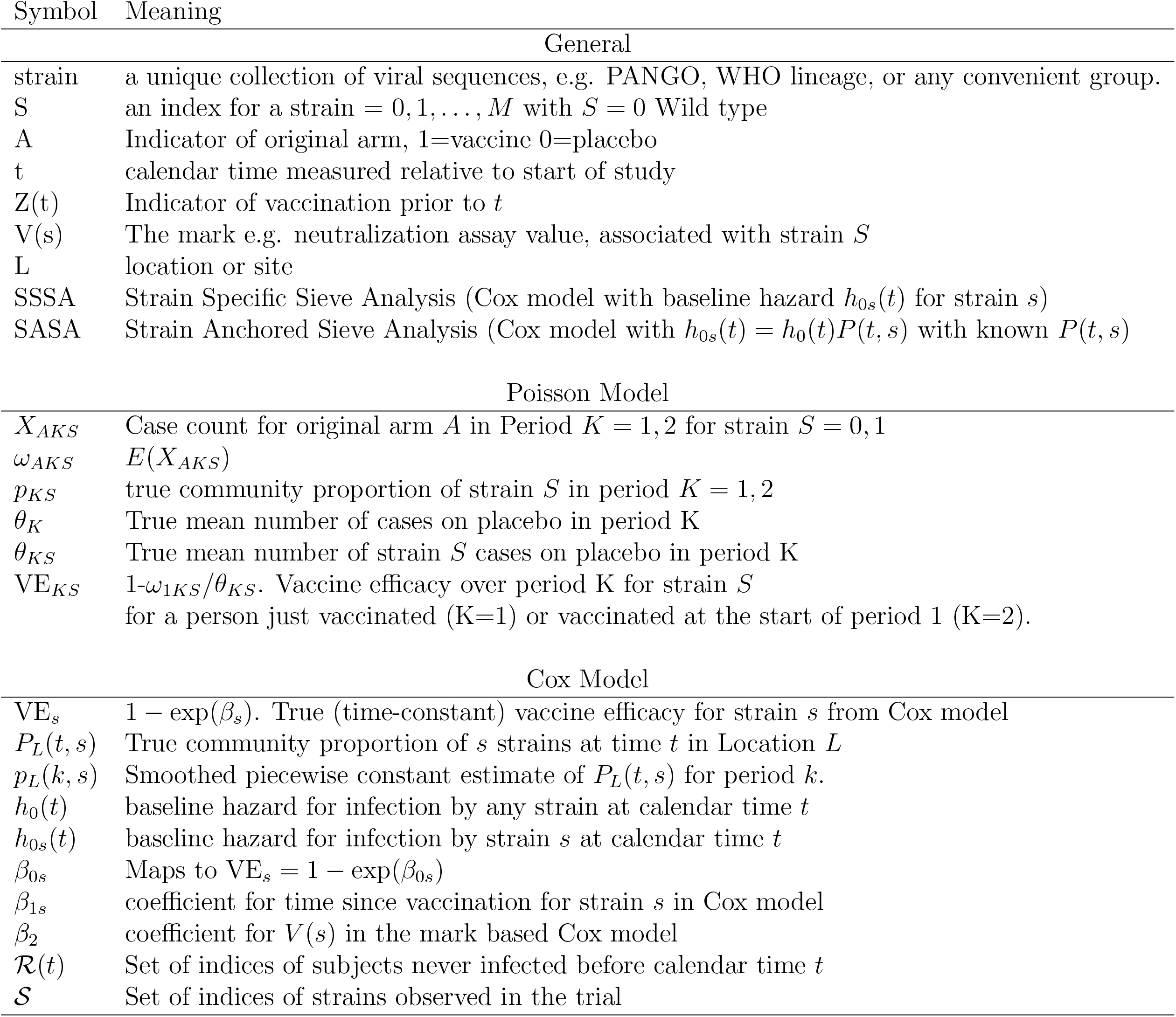
Notation

| Symbol | Meaning |
| --- | --- |
| General |  |
| strain | a unique collection of viral sequences, e.g. PANGO, WHO lineage, or any convenient group. |
| $S$ | an index for a strain $= 0, 1, \dots, M$ with $S = 0$ Wild type |
| $A$ | Indicator of original arm, 1=vaccine 0=placebo |
| $t$ | calendar time measured relative to start of study |
| $Z(t)$ | Indicator of vaccination prior to $t$ |
| $V(s)$ | The mark e.g. neutralization assay value, associated with strain $S$ |
| $L$ | location or site |
| SSSA | Strain Specific Sieve Analysis (Cox model with baseline hazard $h_{0s}(t)$ for strain $s$) |
| SASA | Strain Anchored Sieve Analysis (Cox model with $h_{0s}(t) = h_0(t)P(t, s)$ with known $P(t, s)$) |
| Poisson Model |  |
| $X_{AKS}$ | Case count for original arm $A$ in Period $K = 1, 2$ for strain $S = 0, 1$ |
| $\omega_{AKS}$ | $E(X_{AKS})$ |
| $p_{KS}$ | true community proportion of strain $S$ in period $K = 1, 2$ |
| $\theta_K$ | True mean number of cases on placebo in period $K$ |
| $\theta_{KS}$ | True mean number of strain $S$ cases on placebo in period $K$ |
| $VE_{KS}$ | $1 - \omega_{1KS}/\theta_{KS}$. Vaccine efficacy over period $K$ for strain $S$<br>for a person just vaccinated ($K=1$) or vaccinated at the start of period 1 ($K=2$). |
| Cox Model |  |
| $VE_s$ | $1 - \exp(\beta_s)$. True (time-constant) vaccine efficacy for strain $s$ from Cox model |
| $P_L(t, s)$ | True community proportion of $s$ strains at time $t$ in Location $L$ |
| $p_L(k, s)$ | Smoothed piecewise constant estimate of $P_L(t, s)$ for period $k$. |
| $h_0(t)$ | baseline hazard for infection by any strain at calendar time $t$ |
| $h_{0s}(t)$ | baseline hazard for infection by strain $s$ at calendar time $t$ |
| $\beta_{0s}$ | Maps to $VE_s = 1 - \exp(\beta_{0s})$ |
| $\beta_{1s}$ | coefficient for time since vaccination for strain $s$ in Cox model |
| $\beta_2$ | coefficient for $V(s)$ in the mark based Cox model |
| $\mathcal{R}(t)$ | Set of indices of subjects never infected before calendar time $t$ |
| $\mathcal{S}$ | Set of indices of strains observed in the trial |

**Table 6:** Monte Carlo estimates and (standard deviations) of vaccine efficacy for 100 simulated deferred vaccination trials. Estimates are provided both for strain specific and mark parameterized vaccine efficacy models. Two scenarios are evaluated with VE_0_, VE_1_, VE_2_ = (0.90, 0.90, 0.90) for the top half and with VE_0_, VE_1_, VE_2_ = (0.92,0.78,0.39) for the bottom half. VE is constant over time. The left half fits models with a time constant VE, while the right half fits models with a log-linear decline in VE. The unstarred rows denote correct specification of *p*(*k, s*) while * (**) denote mis-specifications.

|  | Constant VE |  |  | Time-Varying VE |  |  |
| --- | --- | --- | --- | --- | --- | --- |
| | $VE_0$ | $VE_1$ | $VE_2$ | $\beta_{10}$ | $\beta_{11}$ | $\beta_{21}$ |
| TRUTH | 0.90 | 0.90 | 0.90 | 0.00 | 0.00 | 0.00 |
| SSSA | 0.897<br>(0.0364) | 0.885<br>(0.0900) | - | -0.014<br>(0.1483) | 0.001<br>(0.1029) | -0.072<br>(0.5695) |
| SASA | 0.897<br>(0.0356) | 0.890<br>(0.0628) | 0.877<br>(0.124) | -0.016<br>(0.1480) | -0.001<br>(0.0916) | 0.018<br>(0.2600) |
| SASA-MARK | 0.896<br>(0.0349) | 0.896<br>(0.0535) | 0.873<br>(0.1270) |  |  |  |
| SASA* | 0.900<br>(0.0347) | 0.865<br>(0.0797) | 0.849<br>(0.1527) | -0.034<br>(0.1495) | -0.001<br>(0.0905) | 0.044<br>(0.2628) |
| SASA-MARK* | 0.898<br>(0.0341) | 0.885<br>(0.0596) | 0.844<br>(0.1585) |  |  |  |
| SASA** | 0.900<br>(0.0356) | 0.875<br>(0.0580) | 0.909<br>(0.0777) | -0.024<br>(0.1373) | -0.010<br>(0.0958) | 0.037<br>(0.2632) |
| SASA-MARK** | 0.895<br>(0.0353) | 0.904<br>(0.0435) | 0.901<br>(0.0821) |  |  |  |
| TRUTH | 0.92 | 0.78 | 0.39 | 0.00 | 0.00 | 0.00 |
| SSSA | 0.916<br>(0.0317) | 0.754<br>(0.1489) | - | -0.014<br>(0.1815) | -0.003<br>(0.0592) | 0.002<br>(0.0496) |
| SASA | 0.916<br>(0.0311) | 0.767<br>(0.1132) | 0.326<br>(0.3995) | -0.022<br>(0.1708) | -0.001<br>(0.0567) | 0.001<br>(0.0483) |
| SASA-MARK | 0.916<br>(0.0284) | 0.769<br>(0.0853) | 0.320<br>(0.3963) |  |  |  |
| SASA* | 0.918<br>(0.0351) | 0.719<br>(0.1442) | 0.191<br>(0.4930) | -0.036<br>(0.1756) | -0.016<br>(0.0568) | 0.013<br>(0.0482) |
| SASA-MARK* | 0.914<br>(0.0287) | 0.750<br>(0.0951) | 0.217<br>(0.4798) |  |  |  |
| SASA** | 0.921<br>(0.0306) | 0.766<br>(0.0961) | 0.5565<br>(0.2206) | -0.036<br>(0.1542) | -0.016<br>(0.0583) | 0.011<br>(0.0481) |
| SASA-MARK** | 0.911<br>(0.0295) | 0.815<br>(0.0671) | 0.590<br>(0.2234) |  |  |  |

## 7 Discussion

The impact of new strains or variants of SARS-CoV2-2 on vaccine efficacy is of major concern. In this paper we provide methods that permit estimation of strain specific vaccine efficacy after the placebo group is vaccinated. The simplest and most robust is to apply the deferred vaccination methods separately to each strain. However this approach is only viable for strains that occur while there is still a placebo group. The major contribution of this paper is to demonstrate how to use vaccine trial data coupled with strain surveillance data to estimate strain-specific placebo controlled vaccine efficacy profiles for variants that emerge after all are vaccinated. The key requirement is that the distribution of strains for a counterfactual placebo group matches that within the community. This assumption might be violated for several reasons, e.g., the trigger for inputing in the surveillance dataset is different from the case definition of disease in the trial and this changes the distribution of variants, or the surveillance data might draw from a different sublocation than the trial participants and the distribution of variants differs. This assumption can be examined during placebo controlled periods of a trial, but post crossover it cannot be directly examined. In practice we advocate sensitivity analyses where the estimated placebo strain distribution is realistically varied to provide a range of VE estimates.

## Supporting information

GISAIDacknowledgment

## Data Availability

A list of all
submitting and originating
laboratories for all of the sequences used in this study are available
in the supplementary materials.

## Acknowledgements

We would like to thank the GISAID database for their compilation and curation of the full-genome SARS-CoV-2 sequences that were used in this study, and all of the submitting and originating laboratories for submitting their sequenced genomes for public use. A list of all submitting and originating laboratories for all of the sequences used in this study are available in the supplementary materials.

## 8 Appendix

### 8.1 Notation

### 8.2 Supplement: GLM Analysis of Poisson Model

For ease of exposition, we have assumed that *n*_0_ = *n*_1_ = *n* (i.e., that the trial was a 1:1 randomization, and ensuring that there are equal numbers in both arms). For this supplement we relax that assumption, and assume that *γ* = *n*_1_*/n*_0_. Under this generalization, this is a *γ* : 1 randomization and there are on average *γ* individual(s) in arm 1 for each individual in arm 0. Since vaccine efficacies measure proportions of events, we have

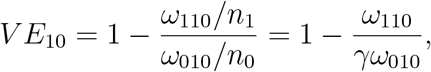

and each individual in arm 0 is scaled to represent *γ* individuals (e.g., in a 2:1 randomization, there are twice as many individuals in the vaccine arm, and each individual in the placebo arm should be multiplied by 2 so the scaled mean for the placebos is twice as large, and the ratio will reduce to the proportions). We use Poisson regression to estimate the parameters of interest. In the generalized linear model in the Poisson family, we model the log of Poisson means as a linear combination of parameters. Let **Y** and *β* be a vector of responses and parameters, with the model of *E*(**Y**) given by

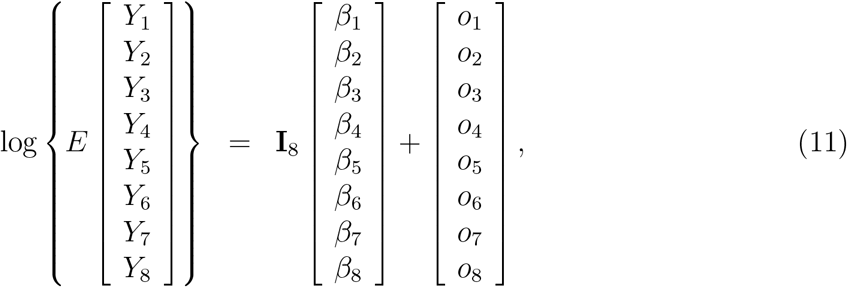

where log(·) and *E*(·) act componentwise (e.g., the log of the vector, is the vector of the log of each component), *I*_8_ is an 8 *×* 8 identity matrix, and *o*_1_, …, *o*_8_ are known constants called offsets. For our application, the responses, parameters and offsets are:

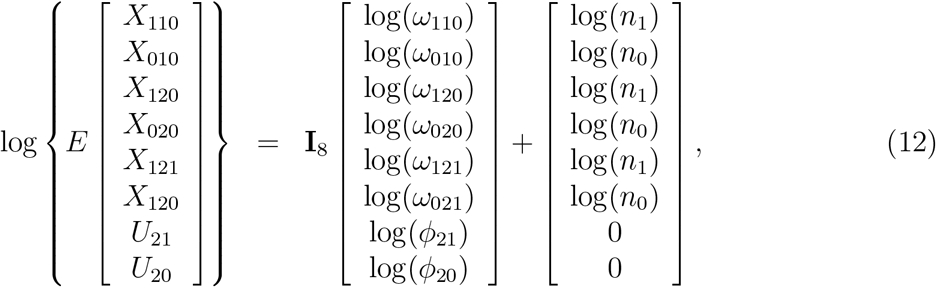

where *U*_2*S*_ are the counts for strain S from the unvaccinated community sample, and the offsets for the first 6 elements ensure that the parameters present rates not counts. Because all of the rates will be later used in ratios of the two arms, we can set *n*_0_ = 1 and *n*_1_ = *γ*. We are interested in the exponentiation of several linear combinations of parameters, specifically,

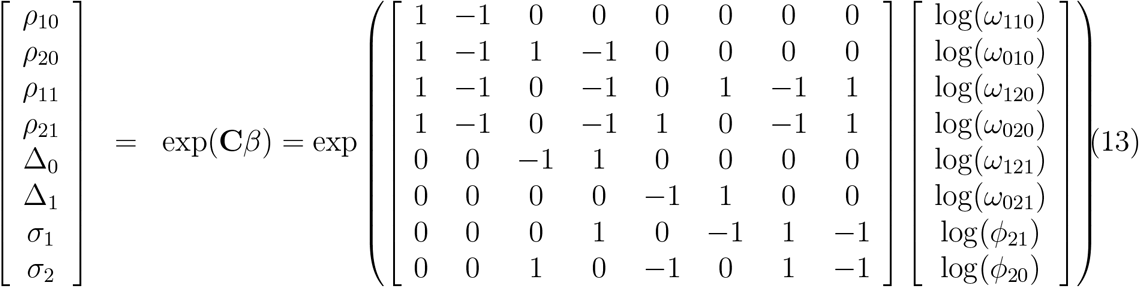

Standard Wald-type confidence intervals can be created assuming that the estimates of log(*ω*_*AKS*_) and log(*φ*_*IS*_) are asymptotically normal. Let 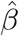 b the vector of those estimates, then we assume that approximately,

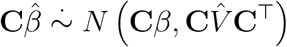

~~~
#
# Computer code to reproduce Table 2
#
 C<- matrix(c(
  1,-1, 0, 0, 0, 0, 0, 0,
  1,-1, 1,-1, 0, 0, 0, 0,
  1,-1, 0,-1, 0, 1,-1, 1,
  1,-1, 0,-1, 1, 0,-1, 1,
  0, 0,-1, 1, 0, 0, 0, 0,
  0, 0, 0, 0,-1, 1, 0, 0,
  0, 0, 0, 1, 0,-1, 1,-1,
  0, 0, 1, 0,-1, 0, 1,-1),
  byrow=TRUE, nrow=8, ncol=8)
 trans<- function(B){ c(1-exp(B[1:4]), exp(B[5:8])) }
simData<-function(parms=c(theta10=1, theta20=1, theta21=1, VE10=.5, VE11=.5, VE20=.5,
 theta10<- parms[“theta10”]
 theta20<- parms[“theta20”]
 theta21<- parms[“theta21”]
 VE10<- parms[“VE10”]
 VE11<- parms[“VE11”]
 VE20<-parms[“VE20”]
 VE21<-parms[“VE21”]
 phi20<-parms[“phi20”]
 phi21<-parms[“phi21”]
 gamma<- parms[“gamma”]
 p20<-theta20/(theta21+theta20)
 p21<-theta21/(theta21+theta20)
Y<-c(
 X110=rpois(1, theta10*(1-VE10)),
 X010=rpois(1, theta10),
 X120=rpois(1, theta20*(1-VE20)),
 X020=rpois(1, theta20*(1-VE10)),
 X121=rpois(1, theta21*(1-VE21)),
 X021=rpois(1, theta21*(1-VE11)),
 U21=rpois(1, phi21),
 U20=rpois(1, phi20))
# MU=true values, so replace N*p21,N*p20 for phi21,phi20
# where N=phi20+phi21
# N<- phi20+phi21
N<-1
MU<- c(theta10*(1-VE10),
               theta10,
               theta20*(1-VE20),
               theta20*(1-VE10),
               theta21*(1-VE21),
               theta21*(1-VE11),
               N*p21,
               N*p20)
n0<- 1
n1<-gamma
Rate<- MU/c(n1,n0,n1,n0,n1,n0,1,1)
untrans.Parms<- C %*% matrix(log(Rate),8,1)
Parms<- trans(untrans.Parms)
names(Parms)<- c(“VE10”,”VE20”,”VE11”,”VE21”,”Delta0”,”Delta1”,”Sigma1”,”Sigma2”)
list(Y=Y, Parms=Parms)
}
glmAnalysis<- function(Y,gamma=1, conf.level=0.95){
   I<- diag(8)
   I1<- I[,1]
   I2<- I[,2]
   I3<- I[,3]
   I4<- I[,4]
   I5<- I[,5]
   I6<- I[,6]
   I7<- I[,7]
   I8<- I[,8]
OFFSET<- log(c(gamma,1,gamma,1,gamma,1,1,1))
gout<- glm(Y∼ -1 + I1+I2+I3+I4+I5+I6+I7+ I8,offset=OFFSET, family=“poisson”)
beta<- matrix(coef(gout),8,1)
V<- vcov(gout)
parms<- C %*% beta
Vparms<- C %*% V %*% t(C)
Za<- qnorm(1-(1-conf.level)/2)
stdErr<- sqrt(diag(Vparms))
parms<- as.vector(parms)
lower<- parms - Za*stdErr
upper<- parms + Za*stdErr
# all null parameter values are zero, log(Ratios)=log(1)=0
p.value<- 2*(1- pnorm(abs(parms/stdErr)))
out<-matrix(NA,8,5)
out[,1]<- parms
out[,2]<- stdErr
out[,3]<- lower
out[,4]<- upper
out[,5]<- p.value
dimnames(out)<-list(c(“logRho10”,”logRho20”,”logRho11”,”logRho21”,”logDelta0”,”logDel
       c(“Estimate”,”StdErr”,”lower CL”,”upper CL”,”two-sided p”))
outVE<- matrix(NA,8,4)
outVE[,1]<- trans(parms)
# for VE, lower and upper switch because transformation is 1-exp(parm)
# for other ratios they do not switch because transformation is exp(parm)
outVE[,2]<- trans(c(upper[1:4],lower[5:8]))
outVE[,3]<- trans(c(lower[1:4],upper[5:8]))
outVE[,4]<- p.value
dimnames(outVE)<-list(c(“VE10”,”VE20”,”VE11”,”VE21”,”Delta0”,”Delta1”,”Sigma1”,”Sigma
          c(“Estimate”,”lower CL”,”upper CL”,”two-sided p”))
list(out=out, outVE=outVE)
}
# Create parameters for all of the simulation Cases
nPHI<- 1e3
THETA2<- 500
parmNames<-c(“theta10”, “theta20”, “theta21”, “VE10”, “VE11”, “VE20”, “VE21”, “phi20”,
PARMS<-matrix(NA,5,length(parmNames),dimnames=list(paste(“Case”,1:5),parmNames))
PARMS[1,]<-c(1000,0.5*THETA2,0.5*THETA2,.9,.9,.9,.9,0.5*nPHI,0.5*nPHI,1)
PARMS[2,]<-c(1000,0.5*THETA2,0.5*THETA2,.9,.8,.8,.6,0.5*nPHI,0.5*nPHI,1)
PARMS[3,]<-c(1000,0.2*THETA2,0.8*THETA2,.9,.9,.9,.9,0.2*nPHI,0.8*nPHI,1)
PARMS[4,]<-c(1000,0.5*THETA2,0.5*THETA2,.9,.9,.9,.9,0.6*nPHI,0.4*nPHI,1)
PARMS[5,]<-c(1000,0.5*THETA2,0.5*THETA2,.9,.9,.9,.9,0.4*nPHI,0.6*nPHI,1)
dosim<-function(nsim=1e2,parms=Parms,…){
  knames<-as.character(1:nsim)
  DimNames<- c(dimnames(g$outVE),list(knames))
  out<- array(NA,dim=c(8,4,nsim),dimnames=DimNames)
  for (k in 1:nsim){
    Yk<- simData(parms)
    Gk<-glmAnalysis(Yk$Y,…) out[,,k]<- Gk$outVE
}
output<- list(out=out, parms=Yk$Parms)
output
}
summarySim<-function(sout,alpha=0.05){
   Table<- matrix(NA,8,7,dimnames=list(dimnames(g$outVE)[[1]],c(“true Value”,”mean”,”std
   Table[,”true Value”]<- sout$parms
   # create 8 X nsim matrix of estimates
   est<- sout$out[,”Estimate”,]
   Table[,”mean”]<- apply(est,1,mean)
   Table[,”std”]<- apply(est,1,sd)
   # create 8 X nsim matrices of lower and upper confidence limits
   lo<- sout$out[,”lower CL”,]
   up<- sout$out[,”upper CL”,]
   nsim<- dim(sout$out)[3]
   # create 8 X nsim
   true parameter matrix trueParm<- matrix(rep(sout$parms,nsim),8,nsim)
   # lower CL too high?
   errloMatrix<- lo > trueParm
   # upper CL too low?
   errhiMatrix<- up < trueParm
   # proportion errors lo=lower too high or hi=upper too low
   Table[,”errlo”]<- apply(errloMatrix,1,mean)
   Table[,”errhi”]<- apply(errhiMatrix,1,mean)
   # proportion with no errors = coverage
   Table[,”coverage”]<- 1 - Table[,”errlo”] - Table[,”errhi”]
   # proportion reject = power
   alphaMatrix<- matrix(alpha,8,nsim)
   pvalue<- sout$out[,”two-sided p”,]
   rejectMatrix<- pvalue <= alphaMatrix
   Table[,”power”]<- apply(rejectMatrix,1,mean)
   Table
}
set.seed(1234)
NSIM<- 1e4
for (k in 1:5){
   set.seed(120*k + k)
   sout<-dosim(nsim=NSIM,parms=PARMS[k,])
   if (k==1){ sumSim<- list(summarySim(sout))
   } else {
            sumSim<- c(sumSim,list(summarySim(sout)))
}
}
getTable2<-function(sumSim){
   tab2<-matrix(NA,10,8,dimnames=list(c(“Case 1”,”“,”Case 2”,”“,”Case 3”,”“,”Case 4”,”“,”
   for (k in 1:5){
     mk<- sumSim[[k]]
     tab2[2*k-1,1:4]<- mk[1:4,”mean”]
     tab2[2*k,1:4]<- mk[1:4,”std”]
     tab2[2*k-1,5:8]<- mk[5:8,”power”]
}
tab2
}
tab2<-getTable2(sumSim)
library(xtable)
print(xtable(tab2,type=“latex”,digits=4),file=“tab2.tex”)
~~~

### 8.3 Example Data Set and SAS code

Here we provide part of a dataset used in the example section. Volunteers can be infected with one of three strains *S* =st=0,1,2 which has mark *V* (*s*) = v =0,1,2. There are 12 months or intervals within which p(k,s) is constant and a single site. Each volunteer generates up to 12 * 3 lines of data for the 12 months times the 3 strains. At the end of the dataset, we provide the code that is used to estimate the SASA with strain specific time-varying vaccine efficacy

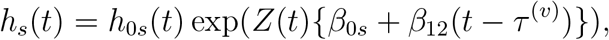

where *t* is calendar time, *Z*(*t*) is 1 following vaccination and 0 otherwise and *τ* ^(*v*)^ is the time of vaccination.

#### 8.3.1 Variable names

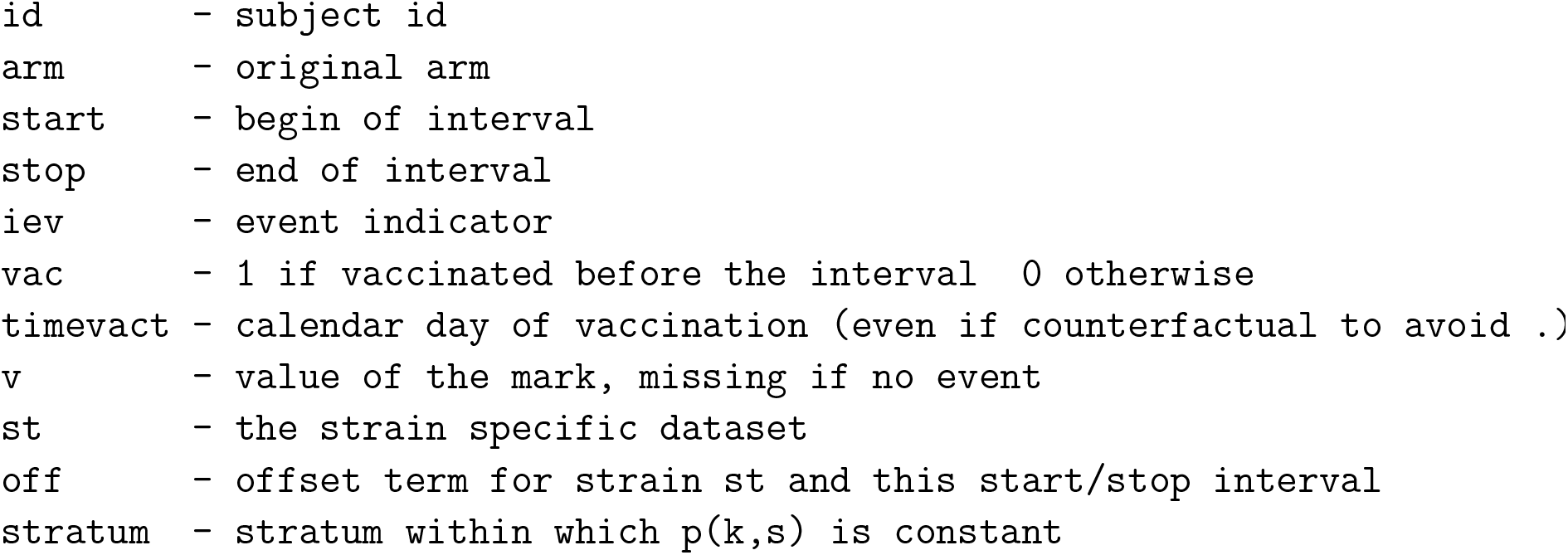

The *p*(*k, s*) distribution is given as

|  |  |  |  |  |  |  |  |  |  |  |  |  |
| --- | --- | --- | --- | --- | --- | --- | --- | --- | --- | --- | --- | --- |
| Stratum | 1 | 2 | 3 | 4 | 5 | 6 | 7 | 8 | 9 | 10 | 11 | 12 |
| Left T | 0 | 1 | 2 | 3 | 4 | 5 | 6 | 7 | 8 | 9 | 10 | 11 |
| Strain | sep | oct | nov | dec | jan | feb | mar | apr | may | jun | jul | aug |
|  | --- | --- | --- | --- | --- | --- | --- | --- | --- | --- | --- | --- |
| 0 -wt | 1.0 | 1.0 | 1.0 | 1.0 | 0.9 | 0.7 | 0.5 | 0.3 | 0.2 | 0.0 | 0.0 | 0.0 |
| 1-alpha | 0.0 | 0.0 | 0.0 | 0.0 | 0.1 | 0.3 | 0.5 | 0.7 | 0.6 | 0.7 | 0.6 | 0.5 |
| 2-delta | 0.0 | 0.0 | 0.0 | 0.0 | 0.0 | 0.0 | 0.0 | 0.0 | 0.2 | 0.3 | 0.4 | 0.5 |

#### 8.3.2 Data Set

ID 1 is a placebo volunteer who enrolls at time 0.0356 and is vaccinated at time 7.000 and is censored at time 12.00. ID 2 is a placebo volunteer who enrolls on 0.4381 and is vaccinated at time 7.000. ID 9 enrolls at time 0.4383 is randomized to vaccine and vaccinated. At time 8.8064 ID 9 becomes a disease case with strain S=1. The offsets were specified as log(p(k,s)) with values *< −* 25 used for p(k,s)=0 to avoid logging zero. By construction, nonzero p(k,s) were always 0.10 or greater.

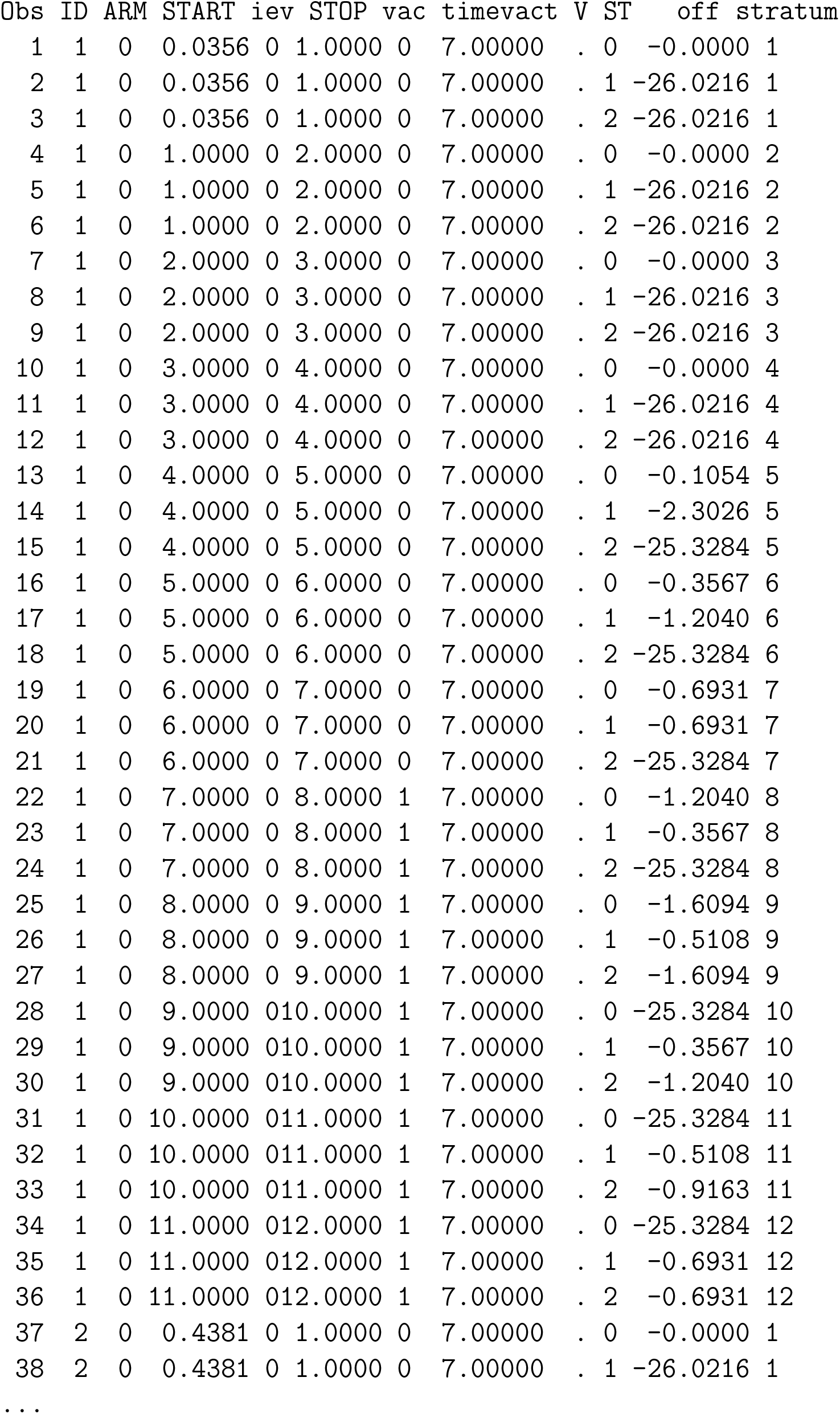
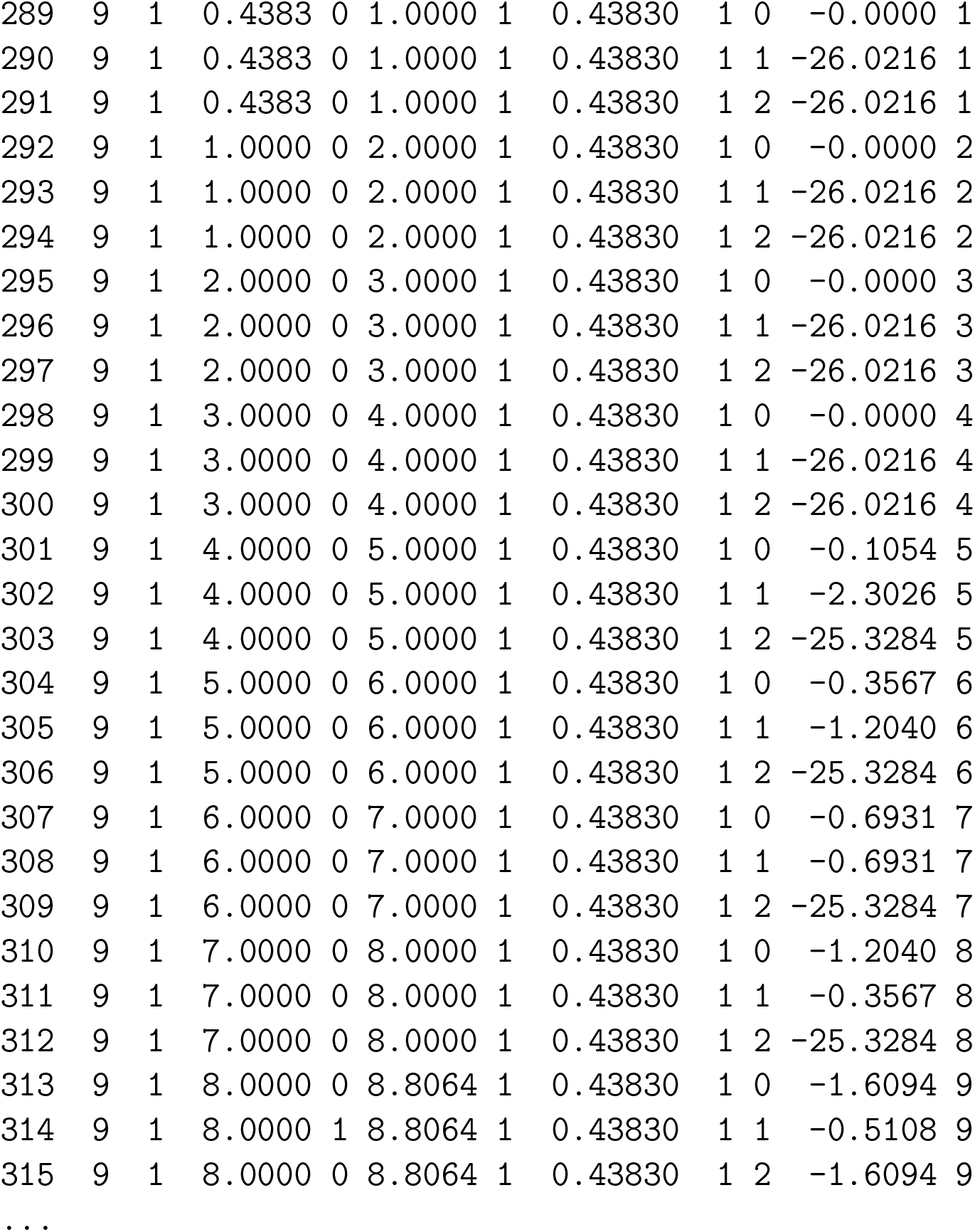

#### 8.3.3 SAS Code for SASA Model

~~~
PROC PHREG DATA=lessbig;
   MODEL (start, stop)*iev(0)= vac0 vactime0 vac1 vactime1 vac2 vactime2/OFFSET=off;
   STRATA stratum;
   vactime0=vac0*(stop-timevact0);
   vactime1=vac1*(stop-timevact1);
   vactime2=vac2*(stop-timevact2);
   IF vac0=0 THEN vactime0=0;
   IF vac1=0 THEN vactime1=0;
   IF vac2=0 THEN vactime2=0;
RUN;
~~~

### 8.4 Additional Simulations

In this section we report results of simulations with the same structure as the simulations reported in Table 3 except the attack rate is much lower, with a pre-crossover placebo attack rate of about 130 and a post-crossover counterfactual placebo attack of about 100. This scenario is meant to approximate the Novavax trial. Results are qualitatively similar to Table 3 but with larger Monte Carlo standard deviations.

## Notes

### Competing Interest Statement

The authors have declared no competing interest.

